# Mind The Gap: Data availability, accessibility, transparency, and credibility during the COVID-19 pandemic, an international comparative appraisal

**DOI:** 10.1101/2022.09.14.22279961

**Authors:** Arianna Rotulo, Elias Kondilis, Thaint Thwe, Sanju Gautam, Özgün Torcu, Maira Vera-Montoya, Sharika Marjan, Md Ismail Gazi, Alifa Syamantha Putri, Rubyath Binte Hasan, Fabia Hannan Mone, Kenya Rodríguez-Castillo, Arifa Tabassum, Zoi Parcharidi, Beverly Sharma, Fahmida Islam, Babatunde Amoo, Lea Lemke, Valentina Gallo

## Abstract

**Background:** Data transparency has played a key role in this pandemic. The aim of this paper is to map COVID-19 data availability and accessibility, and to rate their transparency and credibility in selected countries, by the source of information. This is used to identify knowledge gaps, and to analyse policy implications.

**Methods:** The availability of a number of COVID-19 metrics (incidence, mortality, number of people tested, test positive rate, number of patients hospitalised, number of patients discharged, the proportion of population who received at least one vaccine, the proportion of population fully vaccinated) was ascertained from selected countries for the full population, and for few of stratification variables (age, sex, ethnicity, socio-economic status) and subgroups (residents in nursing homes, inmates, students, healthcare and social workers, and residents in refugee camps).

**Results:** Nine countries were included: Bangladesh, Indonesia, Iran, Nigeria, Turkey, Panama, Greece, the UK, and the Netherlands. All countries reported periodically most of COVID-19 metrics on the total population. Data were more frequently broken down by age, sex, and region than by ethnic group or socio-economic status. Data on COVID-19 is partially available for special groups.

**Conclusions:** This exercise highlighted the importance of a transparent and detailed reporting of COVID-19 related variables. The more data is publicly available the more transparency, accountability, and democratisation of the research process is enabled, allowing a sound evidence-based analysis of the consequences of health policies.

**Funding:** This study was conducted as part of the Summer School “Sustainable Health: designing a new, better normal after COVID-19”. It is a researchers/student collaboration.

## Introduction

As the COVID-19 pandemic is raging across the globe, scientific evidence on transmissibility (1) and the effectiveness of mitigation strategies (2) is accumulating. When translated into health policy, however, this evidence has produced divergent scenarios (3,4). The efficacy of each public health policy could be indirectly evaluated though COVID-19 related data, which have been made publicly available by the majority of the national health authorities, coordinated by the World Health Organisation (WHO) (5), since the very early days of the pandemic (6).

Data transparency has played a key role in this pandemic, facilitating the cross-country comparison of local and national policies, and their evaluation (7). Collective research efforts on data analysis and prediction modelling have fostered dialogue between the scientific community, public health authorities, and policy-makers. However, in many contexts, data availability and transparency have been suboptimal (8,9), a factor that brought a number of negative repercussions in many sectors.

In particular, the breakdown of data reporting by age, sex, region, and ethnic group would help identifying vulnerable groups, which in turn could inform public health strategies and health policies (10). Further stratification, e.g. reporting cases and deaths by occupation, or in specific subgroups (e.g. students, health workers, etc.) would be instrumental to identify patterns of social and health inequalities, and to effectively manage the epidemic at different levels of governance (8) and to ensure political transparency and accountability. To our knowledge, no scientific paper before assessed data availability, accessibility, transparency and credibility internationally, and their related policy implications.

The aim of this paper is to map the availability and transparency of COVID-19 data in selected countries, by source of information, by a number of stratifying variables, and in specific risk groups and to rate their accessibility and credibility. This information is used to identify knowledge gaps, and to analyse policy implications.

## Methods

The Summer School in “Sustainable Health - Designing a new, better normal after COVID-19” took place remotely between the 5^th^ and the 10^th^ of July at Campus Fryslân, University of Groningen. The Summer School attracted a total of 21 students from 14 different countries, from the five continents. All students were postgraduates with a medical/health-related or social science background. During the week, the students were invited to identify COVID-19-related data available in their own countries (either their country of origin or of residence, whose language they were proficient in) and to map their different sources. Each student filled in a shared spreadsheet prepared in advance by three co-authors (AR, EK, and VG). As part of the exercise, students were also invited to rank both the overall accessibility and the credibility of the information. During the last session of the summer school, students were divided into groups and asked present their findings to the whole group.

### Extraction of data: availability and transparency

The extraction tables were designed to be filled in with information from each of the included countries. Information to be collected included the *availability* to the following periodically reported items: i) number of new COVID-19 cases (incidence); ii) number of COVID-19 death (mortality); iii) number of people tested for COVID-19; iv) COVID-19 positive rate (number of those testing positive out of the total number of people tested); v) number of patients hospitalised with COVID-19; vi) number of patients discharged after being hospitalised for COVID-19; vii) proportion of the population who received at least one vaccine; viii) proportion of population fully vaccinated (2/2 or 1/01 at the time, depending on the types of vaccine).

The availability of the information described above was collected, by country, for the full population, and by a number of stratification variables and categories/subgroups. The stratification variables included: 1) age; 2) sex; 3) subnational regions; 4) ethnic background; 5) socio-economic status.

The overall data *transparency* was qualitatively evaluated according to the number of special categories/subgroups data was regularly reported for. These were: a) residents in nursing homes; b) inmates; c) students; d) healthcare and social workers; e) refugees or residents in refugee camps. In addition, the availability of information on the number and size of outbreaks in long-term care facilities, refugee camps, prisons, schools/universities, factories, and nosocomial institutions was also recorded by country.

### Accessibility and credibility of information

By the end of the exercise, a questionnaire was distributed to all participants asking about the accessibility of data in their researched country, and an overall evaluation of data quality and credibility in function of the sources. for *accessibility*, students were asked to rate how difficult it was to find the relevant data from 1 (very easy) to 5 (very difficult). For *credibility*, they were asked to rate how credible they thought data coming from official and unofficial sources were from 1 (not credible at all) to 5 (completely credible). This judgment is based on informal knowledge of the discourse around COVID-19 data availability in their countries, and it has been largely discussed during tutorials.

### Public involvement in the research

This study is a student-teacher collaboration during the online Summer School in Sustainable Health at the University of Groningen. The activity was methodologically led by tutors (AR, EK) and relied on the expertise and contextual knowledge of the students, who contributed to the debate about the importance of data accessibility with examples from their contexts, and who also took an active role in writing this paper.

## Results

A total of nine countries were included in the exercise, with the UK being split into England, Wales, Scotland, and Northern Ireland, resulting in a total of 12 individual country policies. Of these, four (Bangladesh, Indonesia, Iran, Nigeria) were classified as lower-middle income countries; two (Turkey, Panama) as upper-middle income countries; and three (Greece, Netherlands, United Kingdom) as high-income countries, according to the latest World Bank classification(11). For each country, at least one person fluent in the official language of the country and with some public health/health system background, familiarised with the main data repositories and websites of public relevance and was responsible for data searching and extraction.

### Data availability and accessibility

Data availability for the included countries is shown in Figure 1. All countries regularly reported the total number of COVID-19 cases, mortality, testing, hospital admissions, and vaccination from official sources periodically, with few exceptions: in Nigeria the COVID-19 positive rate, the hospitalisation, and the proportion of the population partially vaccinated were not available. In the Netherlands, the number of people discharged after being treated for COVID-19 was not available.

**Figure 1:**
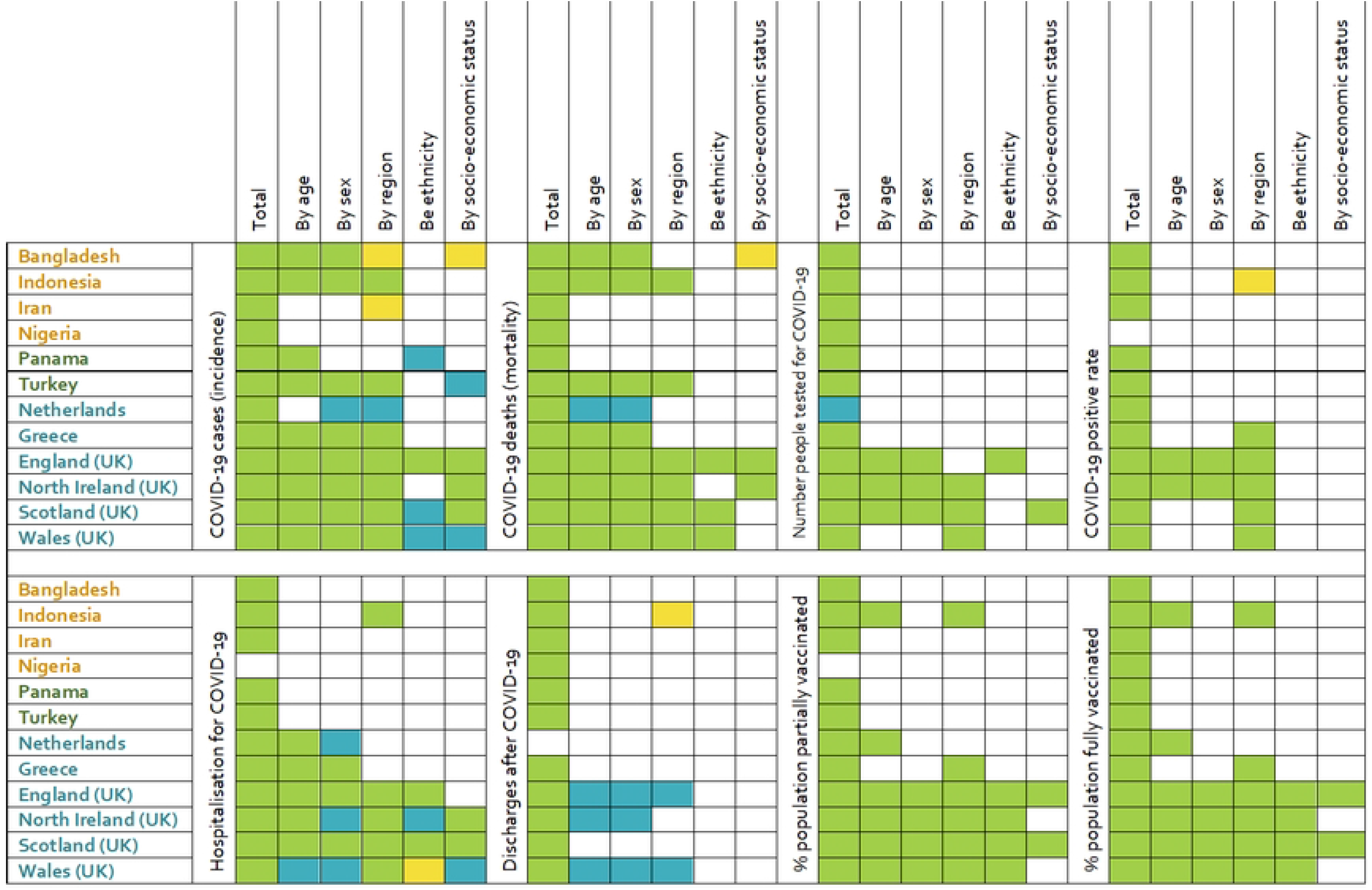
Heatmap illustrating the availability of data on a number of COVID-19 variables in 12 selected countries (divided into lower middle-income in orange; upper middle-income in dark green; and high income in teal) in total, and by a number of stratifying variables. Green: complete data from official source; yellow: incomplete data from official source; blue: data from unofficial source.

Data availability of COVID-19 data per stratification variables (age, sex, region, ethnicity, and socio-economic status) are reported in Figure 1. Overall, data were more frequently broken down by age, sex, and region than by ethnic group or socio-economic status. Variations were observed in terms of disaggregated data in the same income category. The only countries reporting an adequate break-down per stratification variables were the British ones. The Netherlands, Greece, and Turkey reported some break-down by age, sex, and region only for incidence, mortality, and hospitalisation data. However, the information did not always come from official sources. Bangladesh and Indonesia reported some break-down by age, sex, and region only for incidence and mortality data. Iran, Nigeria, and Panama reported little to no broken-down data on all COVID-19 Indicators.

Overall, discharge after COVID-19 resulted to be the category with the least data available by stratification variables. COVID-19 incidence, mortality, and hospitalisation were the variables that more often were presented according to different stratification variables categories. Stratification of data on vaccination was reported only in the four UK countries, and partially in Indonesia, the Netherlands, and Greece.

On average, *data accessibility* was considered difficult: on a scale from 1 (very easy) to 5 (very difficult), the mode was 4 (difficult), rated so by 5 participants (35.7%).

### Transparency of data reporting and credibility

Results of the analysis of data in special sub-groups (residents in nursing homes, inmates, students, healthcare & social workers, and refugees) are reported in Figure 2. Data mainly on COVID-19 incidence and mortality is partially available in a number of countries, while data for the other COVID-19 related variables are more scattered. The country which best reports data according to special categories is Scotland with official/unofficial sources covering most of the fields, particularly among students and health care workers (incidence, mortality, number of tests, positivity rate, vaccine). All UK countries, except for Northern Ireland reported data on vaccination among residents in nursing homes and healthcare and social workers, but data on COVID-19 incidence and mortality among healthcare and social workers is incomplete or coming from unofficial sources in England and Wales. COVID-19 among refugees was officially reported only by Bangladesh (incidence, mortality, and testing); in Greece data on incidence was partially complete, in England, Wales, and Scotland the data was reported by unofficial sources. Data on COVID-19 incidence among inmates was sporadically available from official sources only in Bangladesh, Indonesia, England, Northern Ireland, and Scotland; in Panama and the Netherlands the data was gathered from unofficial sources. Data on COVID-19 related mortality were also available in Indonesia, the Netherlands, England, Scotland, and Wales. Data on vaccination was available only in Indonesia from unofficial sources.

**Figure 2:**
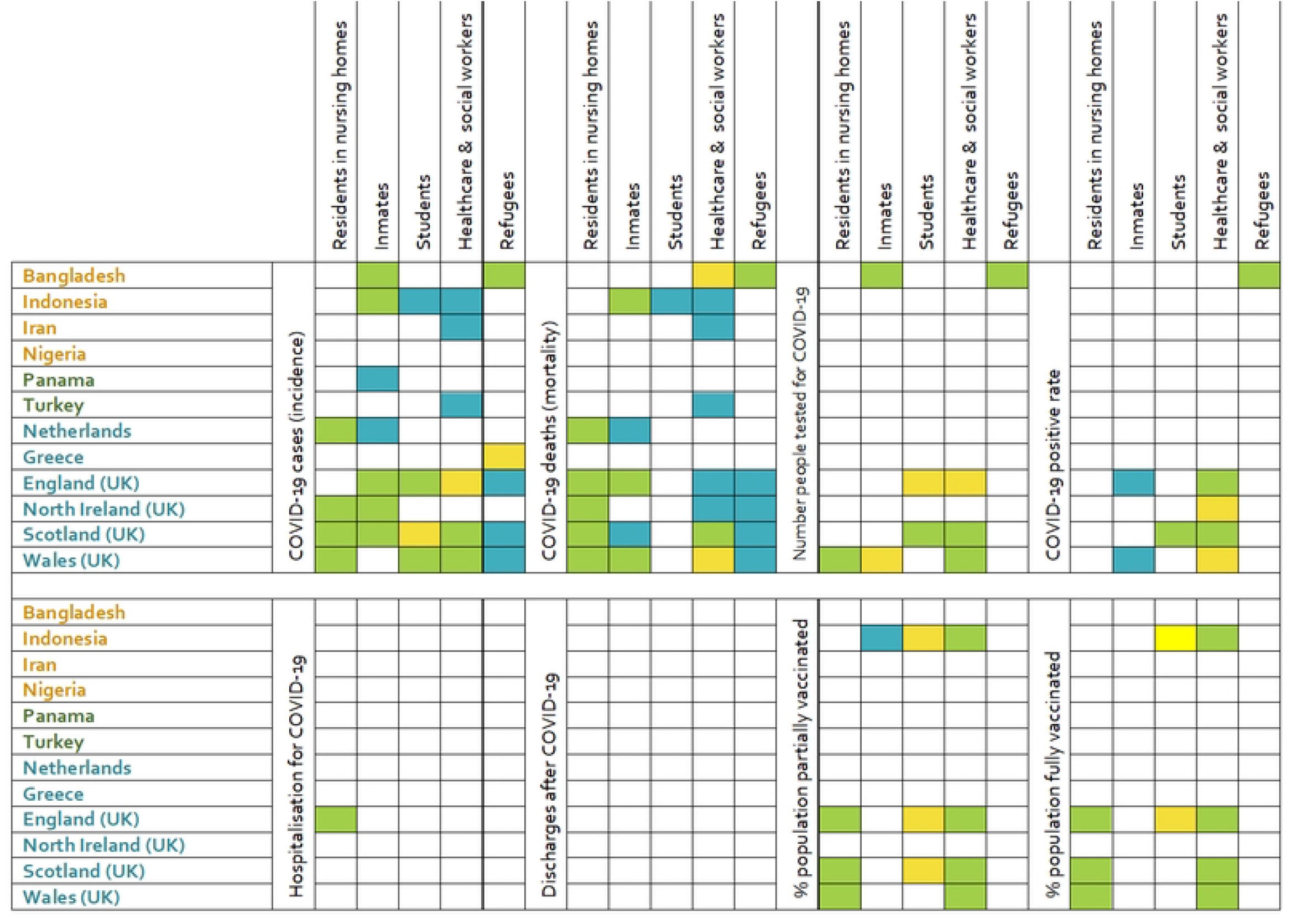
Heatmap illustrating the availability of data in a number of COVID-19 variables in 12 selected countries(divided into lower middle-income in orange; upper middle-income in dark green; and high income in teal) in a number of population sub-groups. Green: complete data from official source; yellow: incomplete data from official source; blue: data from unofficial source

Interestingly, the majority of the authors who extracted the data rated the *credibility* of both official and unofficial sources as high: the mode being in both cases 4 (very credible) rated so by 5 participants (35.7%).

## Discussion

This paper reports a first attempt to appraise in a systematic way COVID-19 related data from a selected number of countries by type of data, stratification variables, and special sub-groups. It prompted a number considerations around the issue of data availability and transparency and the importance of these in pandemic management. Overall, the results suggest an unprecedented effort in collating and making epidemiological data publicly and widely available to the general public from trustworthy sources, despite the fact that such data were considered not always easy to find and access. Varying levels of available budget and infrastructures in high- and low-income countries have not generated significant differences in data availability and accessibility, at least for collated, not stratified data.

Access to stratified data is essential to uncover inequalities in COVID-19 morbidity (10,12–15). Among the included countries, the UK, and – to some extent – Indonesia, had the most accessible data. Refined data availability by regions and Middle-Layer Super Output Areas (MSOA) in England, for example, allowed to explore the relative role of spatial inequalities and of structural factors in explaining the geographical distribution of COVID-19 mortality (14). Availability of data by age and sex allowed to estimate the reduction of life expectancy at birth and lifespan inequalities (16). Data broken down by ethnic group as reported by the Office for National Statistics (ONS) in the UK (17) prompted a parliamentary investigation on why COVID-19 mortality rates were highest among people from Black, Asian, and Minority Ethnic (BAME) groups, with Black males 3.3 times more likely to die compared to their white counterparts (17,18). The investigation resulted in a report (19) suggesting that racism, discrimination, and social inequalities have contributed to the increased risks not only of infection but also of complications and death from COVID-19 among minority ethnic people (18). Importantly, the report emphasised that longstanding inequalities affecting BAME communities in the UK were exacerbated by the conditions under which BAME people live (18). Similar disparities based on ethnicity and migration status were found in other countries such as Sweden (20). The ethnic break-down for vaccine intake is also a crucial piece of information to identify groups whose uptake is suboptimal and to tailor appropriate public health campaigns (21).

When sex-disaggregated data are available, observed inequalities within a country can be appraised in the light of the relative effect of biological factors (22,23) and gender norms (24). Differences between male and female rates of COVID-19 cases and deaths are larger in countries where women experience more discrimination within families and have less access to resources, education, and finance (25). Sex-stratified data in the United States also suggested a different attitude toward vaccine intake between men and women (26). Neglecting sex and gender differences in COVID-19 renders these gender/sex-specific challenges effects unobservable (26). On the other hand, combining such information with data on ethnic background allows an intersectional approach to better understand the relative role of social and biological factors (27).

Data reporting broken down by geographical and demographic strata facilitates international comparison (28) and points out inequalities in varying country contexts (29–31). In the context of vaccination uptake and availability, it can prompt reflections on vaccination equity and the success of the COVAX programme (32).

The transparency of reporting of COVID-19 incidence and mortality in special categories has contributed to a better understanding of the main mechanisms of transmission (33) and the role of inequalities (34), and occupational hazards (35), but has also increased transparency and accountability of health policy decisions. Having observed the very high number of COVID-19-related deaths in nursing homes in England, the UK High Court recently established that the decision – in spring 2020 – to discharge people from hospitals to care homes without mandatory isolation or testing was *irrational* and *unlawful* (36). Data coming from special categories (i.e., prison inmates, people in detention centres and reception centres) can inform the issue of special guidelines for prevention in those contexts (37). Nevertheless, COVID-19 data reporting for these categories remains specifically underreported and therefore understudied (38).

Monitoring of the available COVID-19 data at the international level has been done by several institutions (5,39) and initiatives (40,41). Their work has been extensively used to analyse the rapidly evolving situation (7,42), as well as to estimate international (43,44) and national (45) interventions and policies, and their impact. On the other hand, the unavailability of timely and complete data can elicit misinformation and disinformation among the public, which eventually might hamper the overall health policy enforced (46). The transparent, thorough, and complete report from national authorities has been the necessary first step to allow so.

At the time of writing, the world is living into its third year of COVID-19 pandemic, with an internationally shared sense of grief and fatigue, and uncertainty about the future. It is now more important than ever that the public maintains trust in the institutions (47) and follows government indications to test and receive vaccinations (48–50). Trust in institutions is also likely to induce populations to share crucial information (51) thereby maintaining an effective surveillance system.

Among other things, trust can be enhanced by a transparent and detailed report of available data which increases the accountability of public health authorities (8). Data transparency can also democratise the research effort in fighting the pandemic, ultimately promoting an evidence-based best practice less sensitive to vested interests and political agenda influences.

### Strengths and limitation

This study compares COVID-19 data availability, accessibility, transparency, and credibility in nine resource different and geographically distant countries. Importantly, it maps both official and unofficial sources of information and data access was performed by post-graduate public health/health system professionals who were familiar with the cultural context, the language, and the main reporting sources of each country. Despite the inclusion of more countries would have increased the quality of the cross-sectional comparison, the present data aims at exemplifying the importance of detailed data reporting rather to provide a comprehensive picture.

## Conclusions

In conclusion, this exercise maps a varied combination of COVID-19 related data and their sources. Reported evidence highlighted the importance of a transparent and detailed reporting of COVID-19 related variables by public authorities. The more data is publicly available, the more the research process can benefit from transparency, accountability, and democratisation. This allows a sound evidence-based analysis of the consequences of different health policies. Through this mapping exercise, public health regulators can benchmark how well current information sharing policy is working in different parts of the world. The World Health Organisation can nudge public health authorities, leading the way in improving data sharing.

## Data Availability

Data originates from publicly available sources in different languages and countries

NA

## References

1. Buonanno G, Robotto A, Brizio E, Morawska L, Civra A, Corino F, et al. Link between SARS-CoV-2 emissions and airborne concentrations: Closing the gap in understanding. J Hazard Mater. 2022 Apr 15;428:128279.

2. Czypionka T, Greenhalgh T, Bassler D, Bryant MB. Masks and Face Coverings for the Lay Public : A Narrative Update. Ann Intern Med. 2021 Apr;174(4):511–20.

3. Giritli Nygren K, Olofsson A. Managing the Covid-19 pandemic through individual responsibility: the consequences of a world risk society and enhanced ethopolitics. Journal of Risk Research. 2020 Aug 2;23(7–8):1031–5.

4. Pavlova A, Witt K, Scarth B, Fleming T, Kingi-Uluave D, Sharma V, et al. The Use of Helplines and Telehealth Support in Aotearoa/New Zealand During COVID-19 Pandemic Control Measures: A Mixed-Methods Study. Front Psychiatry. 2021;12:791209.

5. WHO Coronavirus (COVID-19) Dashboard [Internet]. [cited 2021 Jun 14]. Available from: https://covid19.who.int

6. Gallo V, Chiodini P, Bruzzese D, Bhopal R. Age-and sex-adjustment and the COVID-19 pandemic - transformative example from Italy. Int J Epidemiol. 2020 Oct 1;49(5):1730–2.

7. Gallo V, Chiodini P, Bruzzese D, Kondilis E, Howdon D, Mierau J, et al. Comparing the COVID-19 pandemic in space and over time in Europe, using numbers of deaths, crude rates and adjusted mortality trend ratios. Sci Rep. 2021 Aug 12;11(1):16443.

8. Kondilis E, Papamichail D, Gallo V, Benos A. COVID-19 data gaps and lack of transparency undermine pandemic response. J Public Health (Oxf). 2021 Jun 7;43(2):e307–8.

9. Mathieu E. Commit to transparent COVID data until the WHO declares the pandemic is over. Nature. 2022 Feb 22;602(7898):549–549.

10. Bambra C, Riordan R, Ford J, Matthews F. The COVID-19 pandemic and health inequalities. J Epidemiol Community Health. 2020 Nov;74(11):964–8.

11. New World Bank country classifications by income level: 2021-2022 [Internet]. [cited 2022 Feb 28]. Available from: https://blogs.worldbank.org/opendata/new-world-bank-country-classifications-income-level-2021-2022

12. Mena GE, Martinez PP, Mahmud AS, Marquet PA, Buckee CO, Santillana M. Socioeconomic status determines COVID-19 incidence and related mortality in Santiago, Chile. Science. 2021 May 28;372(6545):eabg5298.

13. Mathur R, Rentsch CT, Morton CE, Hulme WJ, Schultze A, MacKenna B, et al. Ethnic differences in SARS-CoV-2 infection and COVID-19-related hospitalisation, intensive care unit admission, and death in 17 million adults in England: an observational cohort study using the OpenSAFELY platform. Lancet. 2021 May 8;397(10286):1711–24.

14. Griffith GJ, Davey Smith G, Manley D, Howe LD, Owen G. Interrogating structural inequalities in COVID-19 mortality in England and Wales. J Epidemiol Community Health. 2021 Dec;75(12):1165–71.

15. Platt L, Warwick R. COVID-19 and Ethnic Inequalities in England and Wales. Fisc Stud. 2020 Jun 3;

16. Aburto JM, Kashyap R, Schöley J, Angus C, Ermisch J, Mills MC, et al. Estimating the burden of the COVID-19 pandemic on mortality, life expectancy and lifespan inequality in England and Wales: a population-level analysis. J Epidemiol Community Health. 2021 Aug;75(8):735–40.

17. Coronavirus (COVID-19) related deaths by ethnic group, England and Wales - Office for National Statistics [Internet]. [cited 2022 Apr 25]. Available from: https://www.ons.gov.uk/peoplepopulationandcommunity/birthsdeathsandmarriages/deaths/articles/coronaviruscovid19relateddeathsbyethnicgroupenglandandwales/2march2020to15may2020

18. Keys C, Nanayakkara G, Onyejekwe C, Sah RK, Wright T. Health Inequalities and Ethnic Vulnerabilities During COVID-19 in the UK: A Reflection on the PHE Reports. Fem Leg Stud. 2021;29(1):107–18.

19. Beyond the Data: Understanding the Impact of COVID-19 on BAME Communities. :69.

20. Bredström A, Mulinari S. Conceptual unclarity about COVID-19 ethnic disparities in Sweden: Implications for public health policy. Health (London). 2022 Feb 13;13634593221074866.

21. Sina-Odunsi AJ. COVID-19 vaccines inequity and hesitancy among African Americans. Clin Epidemiol Glob Health. 2021 Dec;12:100876.

22. Gebhard C, Regitz-Zagrosek V, Neuhauser HK, Morgan R, Klein SL. Impact of sex and gender on COVID-19 outcomes in Europe. Biol Sex Differ. 2020 May 25;11(1):29.

23. Haitao T, Vermunt JV, Abeykoon J, Ghamrawi R, Gunaratne M, Jayachandran M, et al. COVID-19 and Sex Differences: Mechanisms and Biomarkers. Mayo Clin Proc. 2020 Oct;95(10):2189–203.

24. Gausman J, Langer A. Sex and Gender Disparities in the COVID-19 Pandemic. J Womens Health (Larchmt). 2020 Apr;29(4):465–6.

25. Aleksanyan Y, Weinman JP. Women, men and COVID-19. Soc Sci Med. 2022 Feb;294:114698.

26. O’Grady C. Sex and gender missing in COVID-19 data. Science. 2021 Jul 9;373(6551):145.

27. Kopel J, Perisetti A, Roghani A, Aziz M, Gajendran M, Goyal H. Racial and Gender-Based Differences in COVID-19. Front Public Health. 2020;8:418.

28. Mathieu E, Ritchie H, Ortiz-Ospina E, Roser M, Hasell J, Appel C, et al. A global database of COVID-19 vaccinations. Nat Hum Behav. 2021 Jul;5(7):947–53.

29. Duffy C, Newing A, Górska J. Evaluating the Geographical Accessibility and Equity of COVID-19 Vaccination Sites in England. Vaccines (Basel). 2021 Dec 30;10(1):50.

30. McKinnon B, Quach C, Dubé È, Tuong Nguyen C, Zinszer K. Social inequalities in COVID-19 vaccine acceptance and uptake for children and adolescents in Montreal, Canada. Vaccine. 2021 Dec 3;39(49):7140–5.

31. Aktürk Z, Linde K, Hapfelmeier A, Kunisch R, Schneider A. COVID-19 vaccine hesitancy in people with migratory backgrounds: a cross-sectional study among Turkish- and German-speaking citizens in Munich. BMC Infect Dis. 2021 Dec 6;21(1):1214.

32. Bolcato M, Rodriguez D, Feola A, Di Mizio G, Bonsignore A, Ciliberti R, et al. COVID-19 Pandemic and Equal Access to Vaccines. Vaccines (Basel). 2021 May 21;9(6):538.

33. Leibowitz AI, Siedner MJ, Tsai AC, Mohareb AM. Association Between Prison Crowding and COVID-19 Incidence Rates in Massachusetts Prisons, April 2020-January 2021. JAMA Intern Med. 2021 Oct 1;181(10):1315–21.

34. Crispim J de A, Ramos ACV, Berra TZ, Santos MSD, Santos FLD, Alves LS, et al. Impact and trend of COVID-19 in the Brazilian prison system: an ecological study. Cien Saude Colet. 2021 Jan;26(1):169–78.

35. Nafilyan V, Pawelek P, Ayoubkhani D, Rhodes S, Pembrey L, Matz M, et al. Occupation and COVID-19 mortality in England: a national linked data study of 14.3 million adults. Occup Environ Med. 2021 Dec 27;oemed-2021-107818.

36. Booth R, correspondent RBS affairs. Covid care home discharge policy was unlawful, says court. The Guardian [Internet]. 2022 Apr 27 [cited 2022 Apr 28]; Available from: https://www.theguardian.com/world/2022/apr/27/covid-discharging-untested-patients-into-care-homes-was-unlawful-says-court

37. Wang J, Yang W, Pan L, Ji JS, Shen J, Zhao K, et al. Prevention and control of COVID-19 in nursing homes, orphanages, and prisons. Environ Pollut. 2020 Nov;266(Pt 1):115161.

38. Lemasters K, McCauley E, Nowotny K, Brinkley-Rubinstein L. COVID-19 cases and testing in 53 prison systems. Health Justice. 2020 Dec 11;8(1):24.

39. COVID-19 situation update for the EU/EEA, as of 27 April 2022 [Internet]. European Centre for Disease Prevention and Control. [cited 2022 Apr 29]. Available from: https://www.ecdc.europa.eu/en/cases-2019-ncov-eueea

40. Coronavirus Pandemic (COVID-19) - Our World in Data [Internet]. [cited 2022 Apr 29]. Available from: https://ourworldindata.org/coronavirus

41. COVID-19 Disability Rights Monitor [Internet]. [cited 2022 Jul 23]. Available from: https://www.covid-drm.org/data

42. Gallo V, Vineis P, Cancellieri M, Chiodini P, Barker RA, Brayne C, et al. Exploring causality of the association between smoking and Parkinson’s disease. Int J Epidemiol. 2018 Nov 20;

43. Kennedy DS, Vu VK, Ritchie H, Bartlein R, Rothschild O, Bausch DG, et al. COVID-19: Identifying countries with indicators of success in responding to the outbreak. Gates Open Res. 2020;4:62.

44. Braithwaite J, Tran Y, Ellis LA, Westbrook J. The 40 health systems, COVID-19 (40HS, C-19) study. Int J Qual Health Care. 2021 Feb 20;33(1):mzaa113.

45. Spooner F, Abrams JF, Morrissey K, Shaddick G, Batty M, Milton R, et al. A dynamic microsimulation model for epidemics. Soc Sci Med. 2021 Dec;291:114461.

46. Baines D, Elliott R. Defining misinformation, disinformation and malinformation: An urgent need for clarity during the COVID-19 infodemic [Internet]. Department of Economics, University of Birmingham; 2020 Apr [cited 2022 May 8]. Available from: https://econpapers.repec.org/paper/birbirmec/20-06.htm

47. Ayalon L. Trust and Compliance with COVID-19 Preventive Behaviors during the Pandemic. Int J Environ Res Public Health. 2021 Mar 5;18(5):2643.

48. González-Melado FJ, Di Pietro ML. The vaccine against COVID-19 and institutional trust. Enferm Infecc Microbiol Clin (Engl Ed). 2021 Dec;39(10):510–5.

49. Bogart LM, Ojikutu BO, Tyagi K, Klein DJ, Mutchler MG, Dong L, et al. COVID-19 Related Medical Mistrust, Health Impacts, and Potential Vaccine Hesitancy Among Black Americans Living With HIV. J Acquir Immune Defic Syndr. 2021 Feb 1;86(2):200–7.

50. Benin AL, Wisler-Scher DJ, Colson E, Shapiro ED, Holmboe ES. Qualitative analysis of mothers’ decision-making about vaccines for infants: the importance of trust. Pediatrics. 2006 May;117(5):1532–41.

51. Lu L, Liu J, Yuan YC, Burns KS, Lu E, Li D. Source Trust and COVID-19 Information Sharing: The Mediating Roles of Emotions and Beliefs About Sharing. Health Educ Behav. 2021 Apr;48(2):132–9.

